# New implementation of data standards for AI in oncology. Experience from the EuCanImage project

**DOI:** 10.1101/2024.03.15.24303032

**Authors:** Teresa García-Lezana, Maciej Bobowicz, Santiago Frid, Michael Rutherford, Mikel Recuero, Katrine Riklund, Aldar Cabrelles, Marlena Rygusik, Lauren Fromont, Roberto Francischello, Emanuele Neri, Salvador Capella, Arcadi Navarro, Fred Prior, Jonathan Bona, Pilar Nicolas, Martijn P. A. Starmans, Karim Lekadir, Jordi Rambla, EuCanImage Consortium

## Abstract

**Background:** An unprecedented amount of personal health data, with the potential to revolutionise precision medicine, is generated at healthcare institutions worldwide. The exploitation of such data using artificial intelligence relies on the ability to combine heterogeneous, multicentric, multimodal and multiparametric data, as well as thoughtful representation of knowledge and data availability. Despite these possibilities, significant methodological challenges and ethico-legal constraints still impede the real-world implementation of data models.

**Technical details:** The EuCanImage is an international consortium aimed at developing AI algorithms for precision medicine in oncology and enabling secondary use of the data based on necessary ethical approvals. The use of well-defined clinical data standards to allow interoperability was a central element within the initiative. The consortium is focused on three different cancer types and addresses seven unmet clinical needs. We have conceived and implemented an innovative process to capture clinical data from hospitals, transform it into the newly developed EuCanImage data models and then store the standardised data in permanent repositories. This new workflow combines recognized software (REDCap for data capture), data standards (FHIR for data structuring) and an existing repository (EGA for permanent data storage and sharing), with newly developed custom tools for data transformation and quality control purposes (ETL pipeline, QC scripts) to complement the gaps.

**Conclusion:** This article synthesises our experience and procedures for healthcare data interoperability, standardisation and reproducibility.

## BACKGROUND

Artificial intelligence (AI) for oncology is an exponentially growing field[1] built over large amounts of patient-related data. The volume and depth of personal health data necessary to support precision medicine is unprecedented, and the integration and analysis of such heterogeneous data types require a thoughtfully structured representation of knowledge[2,3]. Besides, data needs to be shared across diverse institutions and even across multiple nations, increasing the complexity of data integration and flows. FAIR principles (Findability, Accessibility, Interoperability and Reusability) are the international reference that defines the best practices for data sharing. ‘Findability’ entails the automatic discovery of datasets and services. ‘Accessibility’ references the retrieval of the data, possibly including an approval process. ’Interoperability’ refers to the use of standards for data integration. Finally, ‘Reusability’ implies an adequate description of the data (metadata) to optimise their use[4]. Adherence to the FAIR principles is imperative, and its application in health science and oncology is crucial for developing data sharing platforms[5].

Personal health data includes a wide range of different data types, among others, demographic characteristics, patient’s symptoms, diagnoses, laboratory results, medications, imaging data and genomics. Generating and collecting these data types involves a multitude of different technological platforms (e.g. different vendors) and their storage in a wide range of data formats and information systems used by the different healthcare facilities, contributing to increased data diversity[6]. In fact, healthcare data has been shown to be more heterogeneous than other types of research data[7]. This high data complexity makes interoperability of healthcare information a significant hurdle in the development of AI models[8,9]. The barrier is even more prominent in oncological observational research since cancer diagnoses require a set of attributes usually registered separately (imaging, histology, topology, grade, stage, and biomarkers), and the complex patient trajectory involves personalised treatment regimens[9].

Data needs to be harmonised at three levels to address the interoperability challenge: technical, syntactic and semantic. On one hand, requirements in technical interoperability facilitate basic data exchange conventions (file formats), and on the other hand, syntactic and semantic interoperability define data structure and the use of ontologies for unambiguous representation of medical concepts, respectively[10]. Across the different healthcare ecosystems, equivalent information can be represented in diverse ways. The use of standards that can be universally interpreted, both human and machine-readable, facilitates harmonisation efforts. The structured exchange of health-related data is supported by international standards, such as the Fast Healthcare Interoperability Resources (FHIR) specification developed by Health Level Seven (HL7®) international. FHIR defines the structure of medical data in modular components called “Resources”[11]. It is envisioned that the FHIR framework can become critical for implementing AI technologies in the health sector, just as Digital Imaging and Communication in Medicine (DICOM) or Picture Archiving and Communication System (PACS) for imaging data[6].

The EuCanImage initiative is a European Council Research and Innovation Action funded research project that comprises multidisciplinary teams with the overall aim of building a data sharing platform to be filled with over 20,000 cancer cases and AI models integrating imaging, clinical and phenotypic data from five different EU countries to improve cancer patients’ outcomes. Briefly, the EuCanImage platform integrates established data infrastructures: Collective Minds Radiology (CMRAD) platform for collaborative image annotation, the Eurobioimaging for image storage, the European Genome-phenome Archive (EGA) for clinical and phenotypic data storage and the Open EBench platform for AI algorithm benchmarking[12]. In this report, we synthesise our experiences as an overview of the methods and challenges we identified while working towards the standardisation and interoperability of health data (clinical data) and implementing a data model for AI in large-scale oncology research.

## DATA COLLECTION

### Purpose for data collection and data description

EuCanImage is a complex project centred around addressing seven key unmet clinical needs in cancer imaging[12–14]. A multidisciplinary team of different speciality physicians (radiologists, oncologists, radiotherapists, surgical oncologists, pathologists), sociologists, psychologists, AI developers, data scientists, Small and Medium Enterprises (SMEs), imaging and oncology research associations, as well as patients’ organisations, collaborated to identify and refine core research questions. The consortium pinpointed the most urgent topics in liver, colon and breast cancer and designed seven clinical use cases to respond to each specific clinical need. More specifically, we concentrated our efforts on one use case on hepatocellular carcinoma addressing the detection of indeterminate small lesions, three on colorectal cancer: one aimed to identify liver metastasis from pre and post-operative CT in colorectal cancer patients, and two on rectal cancer to a) identify lymph node metastasis in contrast-enhanced rectal MRI and b) predict the response to neoadjuvant treatment and three on breast cancer to a) identify patients likely to achieve pathological complete response to de-escalate neoadjuvant systemic therapy based on the single point, pre-treatment contrast-enhanced MRI, b) automatically differentiate benign and malignant lesions in screening mammograms and c) distinguish molecular subtypes of breast cancer based on digital mammograms. Our ambition is to integrate clinical, pathological and genetic data (non-imaging data) and radiological images (imaging data) to build algorithms going beyond standard practice, allowing personalised approaches informed by the best quality data. The initial effort to obtain such high-quality data was dedicated to defining clinical consensus and requirements for the use cases with specifications of clinical data variables. The mentioned factors necessitate a comprehensive data model incorporating multifactorial inputs from multiple data sources. In EuCanImage, data is submitted from six university hospitals in Italy, Lithuania, Poland, Spain (two sites), and Sweden, national registries and two research institutions from the Netherlands. Each of the centres uses its own imaging infrastructure and PACS as well as electronic or paper health records that include demographic, clinical, pathological and phenotyping information recorded in Health Information Systems.

Regarding the clinical data defined for each use, some common variables exist for all use cases: patient ID, biological sex, age at diagnosis, diagnosis, and pathology (ICD-O-3 codes). On the other hand, there are use case-specific variables such as the hormone receptor status, HER2 mutational status, Ki67 status for breast cancer, or information on specific chemotherapy agents with dosing regimens. The dialogue between physicians and AI developers on clinically relevant variables that can be meaningfully incorporated in AI algorithms, with the GDPR-compliant data minimisation principle, led to the final set of defined variables (Figure 1).

**Figure 1.**
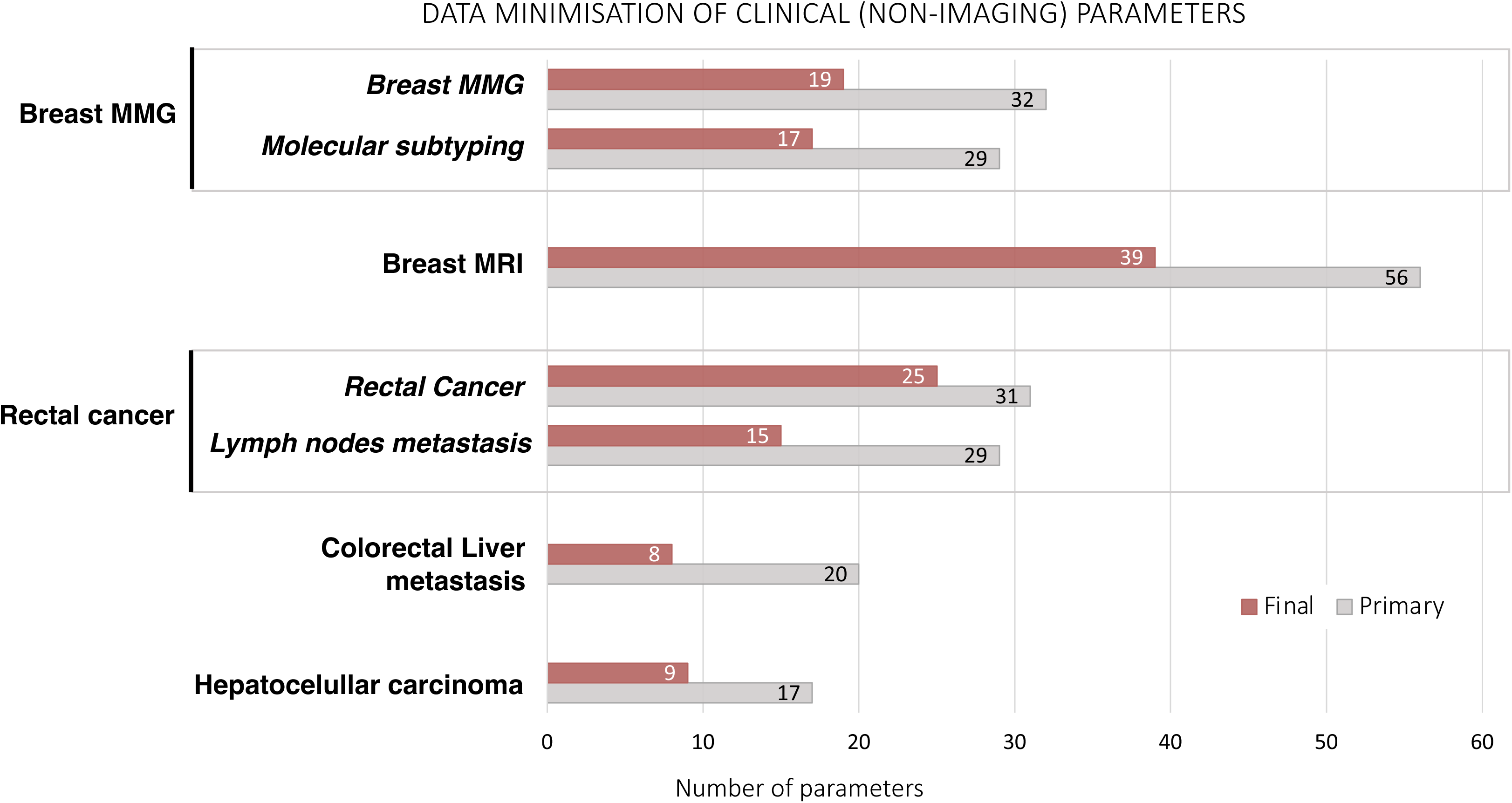
Data minimisation of clinical (non-imaging) parameters

The selection of the clinical variables was a complex and lengthy procedure starting at the beginning of the EuCanImage project with the creation of the Clinical Working Group including clinical representatives from all participating clinical centres. Each clinical site delegated a broad representation of specialists: radiologists, pathologists, clinical oncologists, surgical oncologists, radiotherapy specialists and data managers. In general, the Clinical Working Group meetings were attended by 10-20 doctors every two weeks for the first year of the project. After general concepts of the use cases were established, the Clinical Working Group was divided into organ-related meetings (breast, colorectal and liver subgroups) integrating different specialists per use case and centre. These organ-related specialised groups met every two weeks for the next six months to develop the final list of clinico-pathological variables using the Delphi consensus methodology. The final variables are deemed to provide both the ground truth and the clinical data with additional value for deep learning (DL) modelling.

The final number of clinical variables used as ground truth or additional input parameters for DL, varies between 8 and 39 variables per use case. This clinical information will be used along with the information extracted from radiological images to build next generation deep learning models combining both the clinical and the imaging information simultaneously. Three levels of data provision were defined: minimal, mandatory and recommended. The minimal set contains essential information from the pathology assessment of specimens, e.g., cancer vs. other findings and the presence of complete pathological response vs. partial or no response. It allows the assembly of standard-level algorithms primarily using imaging information as input with pathology information as ground truth. The mandatory set contains important enriching information. These are all variables that should be included as input together with cancer images for more advanced and complex algorithms. Finally, the recommended set addresses additional clinical data points (e.g., risk factors for breast cancer) and phenotyping information (PAM50 results) available only from selected centres but with adequate numbers of patients for AI research. This recommended set would create a very interesting and promising asset in the project repository for future research that is otherwise not readily available from other data repositories.

Next, for each clinical variable, we defined comprehensive and detailed value sets to standardise concept representation and link the terms with ontological codes, ensuring unequivocal understanding. It allows good description of cohorts and, at the same time, prevents very fine-grained stratification of data with limited instances and unbalanced distribution in some of the cohorts. This consensus approach represents a compromise between the need for a precise representation of the clinical range of disease presentations and the goals of data clarity and homogeneity.

## DATA CURATION

### Analysis of semantic interoperability and health standards

Semantic interoperability represents a remarkable challenge for medical research. Data captured through health information systems are usually stored in locally-modelled clinical repositories, mostly in non-structured ways, thus hindering cross-national data source integration and translational research. Health information standards play a crucial role in defining the structure and meaning of clinical information so that different systems can unequivocally interpret it. However, there is no single standard that solves every need in the biomedical field; rather, there are different standards that complement or compete with one another. This includes standardised vocabularies and classifications and also health information standards. Examples of vocabularies include the Systematized Nomenclature of Medicine – Clinical Terms (SNOMED CT)[15] or International Classification of Diseases (ICD)[16], Logical Observation Identifiers Names and Codes (LOINC)[17], OHDSI Standardised Vocabularies, and International Cancer Genome Consortium[18] -Accelerating Research in Genomic Oncology (ICGC ARGO[19]). Examples of health information standards include HL7 FHIR[20] and open Electronic Health Records (openEHR)[21].

Vocabularies and classifications represent concepts that pertain to the biomedical domain in a standard fashion[22], although they require a common structure that provides the syntactic interoperability required to achieve semantic interoperability. Common Data Models (CDMs) serve as representations of collected data aimed at facilitating the exchange, pooling, sharing, or storing data from multiple sources and can provide this common structure[23]. Health information standards also provide a syntactic base to allow the formal representation of the structure of clinical information and its meaning.

FHIR was introduced in 2011 by the standard-developing organisation HL7[20]. The information within FHIR is organised in basic building blocks named Resources. Those blocks define the structure of the contained information. Although it is widely used in health informatics, its uptake in research environments is less prevalent[24]. Most studies using FHIR in health research focus on clinical research (including clinical trials), and just about 12% are oncology-related[25]. In these studies, FHIR has been mainly used for standardisation and data capture and, to a lesser extent, for data analysis[25].

Observational Medical Outcomes Partnership (OMOP) enables the systematic analysis of disparate observational databases through a common data model and a closed dictionary of terminologies, vocabularies, and coding schemes. Several authors consider it an adequate data model for sharing data in electronic health record (EHR)-based longitudinal studies[23,26].

ICGC ARGO[19] is an initiative that provides a fixed schema for creating 15 clinical tables oriented to genomic oncology research, thus oriented for addressing cancer-specific issues in the representation of clinical data. Unlike other CDMs and health information standards, its use of standardised vocabularies and classifications is limited.

### Data Model Design

Data standardisation within the project is required to a) support content organisation and subsequent development of AI algorithms, b) facilitate interoperability and c) the secondary use of the data (ie: data distribution under request in a repository). Both OMOP and FHIR are widely adopted standards in clinical settings; however, they were conceived to serve different purposes. OMOP is more oriented toward clinical data representation (structure and content), and FHIR is more focused on healthcare data exchange. After thoroughly evaluating various CDM alternatives, we decided to use FHIR due to its wide adoption, flexibility and suitability for real-world data exchange. More importantly, and the key aspect we considered in selecting FHIR over OMOP was its appropriateness for permanent data storage and long-term data sharing through the repository.

As previously outlined, clinical elements necessary for each hypothesis were established by domain experts and interdisciplinary teams, including clinicians and AI developers, who considered different key data aspects. Some key considerations for variable selection were characterisation of the target population, clinical endpoints (pathological hallmarks, disease behaviour, treatment response and the patient prognosis), type of outcome (binary, continuous, time to event), adequate ground truth, minimal amount of data principle and the availability of specific variables at data sources. For the project, data to cover the seven use cases were arranged in five different data schemas with single schemas used by multiple use cases. As a general overview, the highest level components of the FHIR model are the Resources, which contain hierarchical sub-layers of descriptive elements for more detailed data classification. The content and format of a Resource have controlled properties, meaning that the different data elements and data types need to adhere to specific requirements. To design the data architecture needed for each EuCanImage use case, the following FHIR resources were identified as relevant: Patient, Condition, Observation, Procedure, Medication Administration and Diagnostic report (Figure 2A). When choosing the most suitable resource for each selected variable, the resource’s constraints were considered. For example, the classification of patients into case (cancer) or control (benign lesion) groups could be interpreted as part of the Condition Resource and information about a diagnosis can be stored in an Observation Resource, capturing the results of tests (mammogram). In this example, we considered it within the Condition Resource despite also including benign cases to associate it with age at diagnosis. Once the variables were assigned to a Resource, they were mapped to the appropriate FHIR element. In cases where the clinical variables could be assigned to more than one suitable profile (ex: Histological type), simplicity criteria were applied to minimise the number of Resources used. Many data elements within the FHIR Resources require coded values. Some are fixed values defined by the FHIR specification, but others require external ontologies. As a general rule, HL7/FHIR terminology was used in a few established fields, specifically status profiles. SNOMED was the preferred terminology for general clinical concepts, ICD-O3 for histology, LOINC for some test observations and RxNORM for medication. We used NCIT when the concept did not exist in previous ontologies (Figure 2B). The summary of the different stages we followed to conceptualise the data model is described in Figure 3.

**Figure 2.**
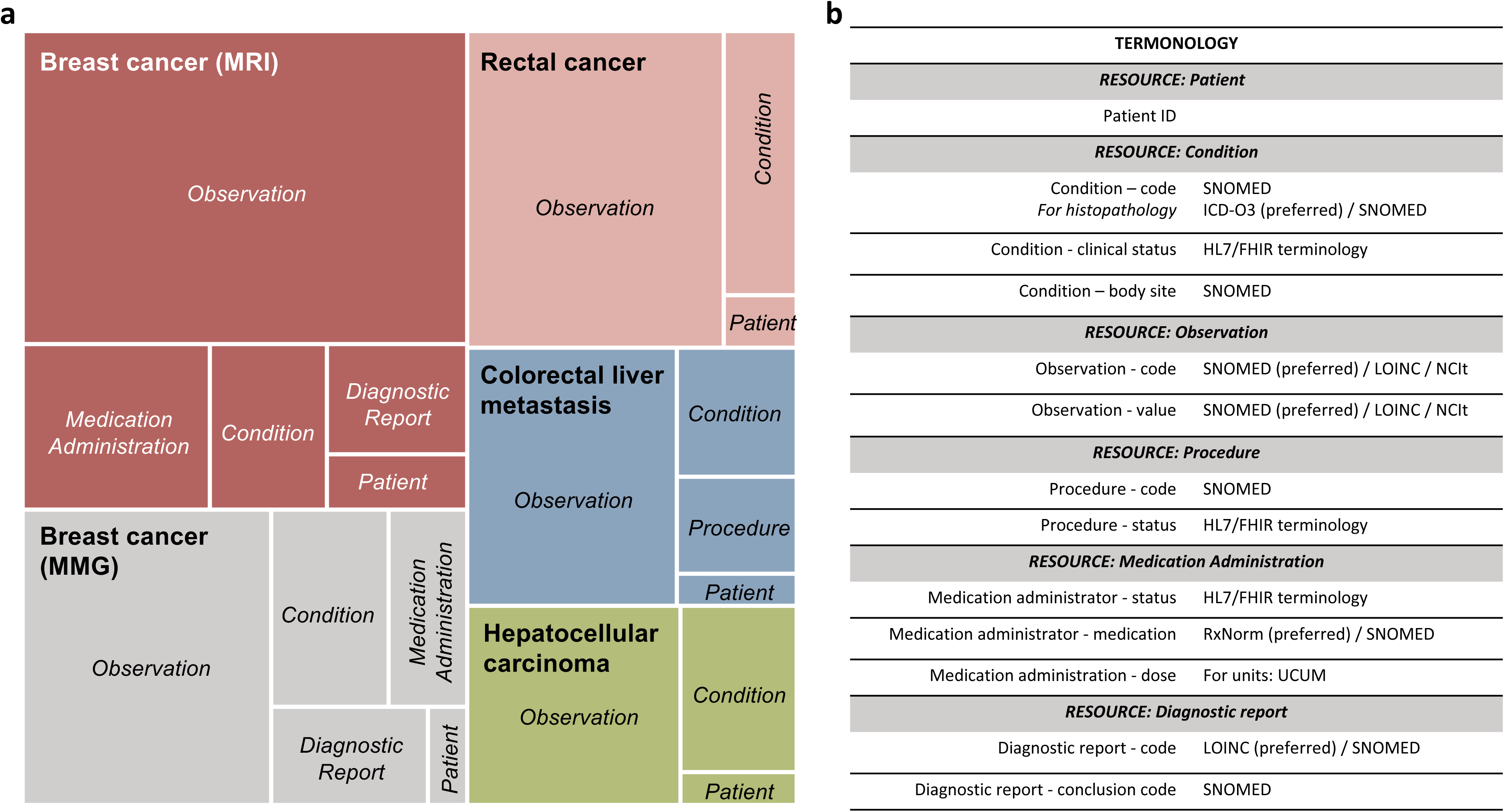
**A)** Representation of the proportion of FHIR resources needed for each use-case **B)** Ontologies used in each resource

**Figure 3.**
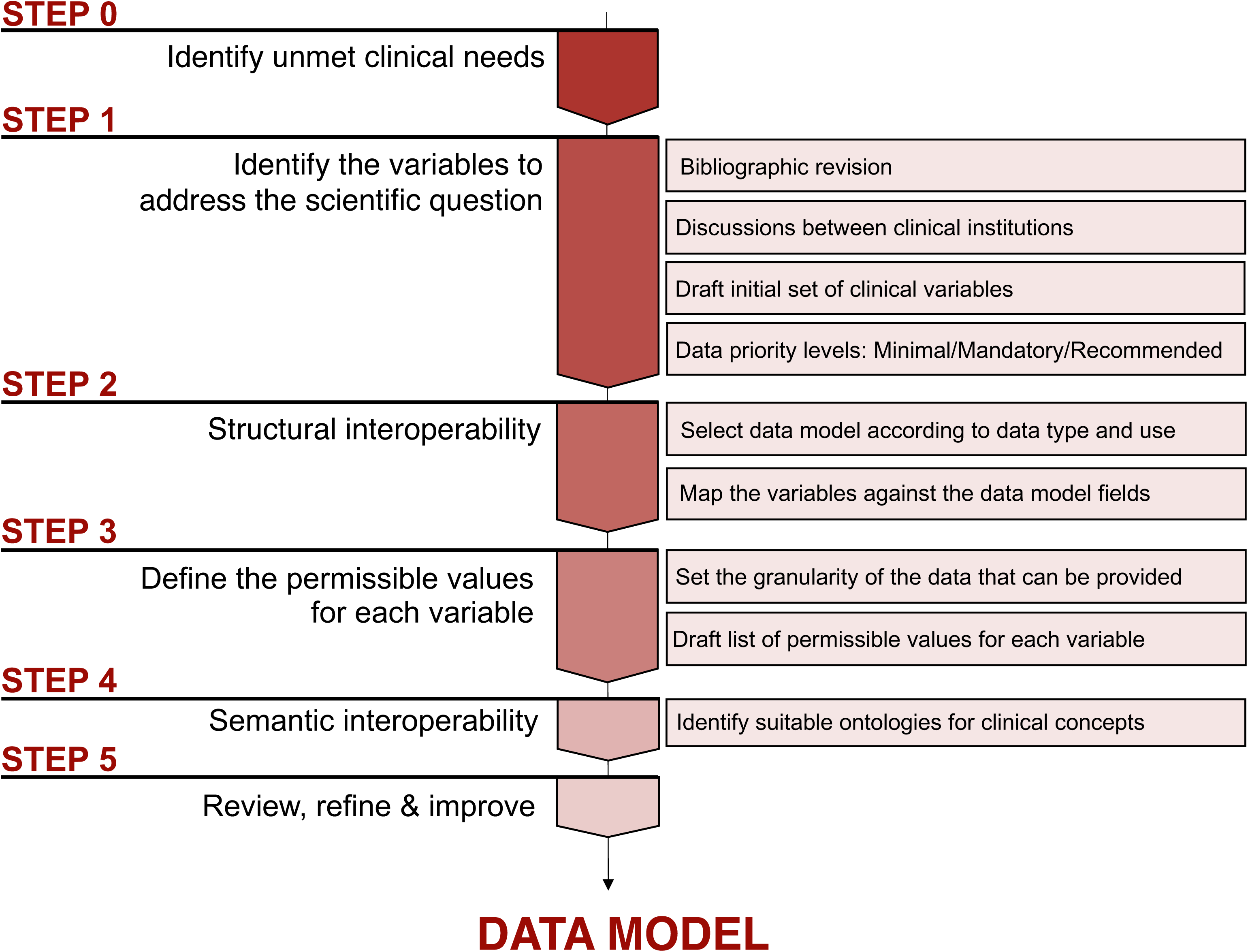
Description of the different steps followed to conceptualise the data model

It is essential to point out that the project presented some particular needs that were not fully represented by FHIR (and standards ontologies), requiring alternative solutions to overcome limitations. The gaps identified relate to the fact that FHIR was designed to support interoperability and data exchange in healthcare rather than specifically focusing on research needs. The primary limitations we faced were a) the need to represent concepts without available standard terminology, b) variables not structured as in healthcare practice, c) the representation of dates to comply with the de-identification of personal data, d) the representation of not provided (missing) information, e) the implementation of the model without a FHIR server.

For each clinical variable, we defined the limited set of permissible values (value set) that this variable can adopt. Some of these value sets needed to include ambiguous concepts for simplicity and data harmonisation reasons. An example is the term ‘other’, which is required to group less frequent or more irrelevant values. Those terms, isolated from additional context, posed a challenge for interoperability. We used SNOMED post-coordinated expressions to build more specific clinical ideas by combining relevant terms with compositional grammar. Another challenge posed was concepts that are not used in healthcare but are essential to contextualise the specific use cases for research purposes and which are not captured by standard terminologies. Some examples are the variable ‘breast cancer subtype-by proxy’ to group breast cancer patients according to hormone receptor, Ki67 and HER2 expression levels or ‘time interval between the end of the neoadjuvant treatment and surgery’. Additional difficulties include tumour grading systems such as the modified Ryan Scheme for Tumor Regression Grade, Miller and Payne’s Tumor Regression Grade, Residual Cancer Burden class or grading of DCIS. Our approach was using the specific grading scales in NCIT, if available, or using generic grading scales in SNOMED (ex: grade 1 on a scale of 1 to 5), despite the fact it could affect interoperability.

Some variables were characterised under the Medication Administration Resource that presented significant difficulties in creating the representations needed for the project. For example, to detail the chemotherapy dosage, we required the total number of chemotherapy cycles, dose (amount of medication per dose) and the accumulation dose within the same ‘Medication administration’ entry. However, that Resource is designed to collect only a single entry.

In compliance with GDPR, personal data pseudonymization entails the removal of indirect data identifiers, such as dates. The collection of exact dates was replaced by the collection time intervals (months, weeks, etc.). Most FHIR Resources allow time periods as valid data types, however the Resource: Medication Administration, only allows dates (*ddmmyy*). To fulfil this FHIR restriction we recodified the time periods into arbitrary dates starting on January 1st of 1970 to mimic the Epoch Unix system, with the end date calculated based on the collected time interval and starting date in mind.

## TECHNICAL IMPLEMENTATION OF DATA STANDARDS

Transforming and loading ‘raw’ data from various hospital data systems into the newly developed data schema proved to be challenging and labour-intensive. The harmonisation efforts required by the different participating institutions and different use cases varied significantly depending on the existing resources at the sites. While some centres housed structured repositories with variables linked to standard terminologies that required minimal mapping and transformation efforts, others mapped local concepts with the project schema manually. This effort was performed by trained site personnel who understood the clinical concepts in both English and the centre’s local language. Online support was provided by the EGA when needed.

The FHIR implementation format has a hierarchical architecture, that, while having many advantages to encode the relationships between the variables and facilitating data storage, supposes an additional barrier for data providers given that most of the required information was not structured data inside their health records. To minimise the need to re-encode and simplify the data capture process, we created electronic case-report forms (eCRF) with REDCap. REDCap is a secure web application that supports data capture primarily for research studies[27,28]. This software allows the custom design of data entry forms and data collection workflows. It features a user-friendly interface for designing the forms, field validation, custom logic patterns, calculated fields, data import/export options, data quality control and role-based user access. Additionally, it offers a set of APIs for integration with other platforms[29]. REDCap was deployed at the European Genome-phenome Archive (EGA) to design and manage data entry forms for clinical data collection within the consortium. Different data entry forms were conceived to support each of the five different data schemas.

Data from hospitals can be imported to the EuCanImage REDCap database following two paths: 1) by directly filling the online forms or 2) by entering data into CSV files complying with specific REDCap format requirements and uploading the files into REDCap.

Patient IDs were previously pseudo-anonymised at the hospitals, and only hashed patient IDs (EuCanImage ID) were introduced in the platform. Consequently, all related clinical data from the different institutions merged into a single harmonised database for each use case. Once harmonised data was in the database, quality control checks were performed. All data was then exported from REDCap as a CSV file for conversion into FHIR-compliant files.

To implement the FHIR model, we created each Resource using individual persistent identifiers with Uniform Resource Names (URN), more specifically with Universally Unique Identifiers (UUID). These identifiers were generated for each patient, resource and bundle. In FHIR, a bundle is a way to gather all the Resources belonging to a single patient. In our case, a bundle is generated from a single row of the exported CSV files.

For the subsequent data standardisation stage, we built Extract Transform Load (ETL) pipelines to transform the output CSV files into JSON files compliant with the FHIR schema. To automate this process, we used Python 3.11, and followed a FHIR 4.3 schema. The python scripts are available on Github (see ‘Availability of supporting source code and requirements’ section), with the additional use of external validators, such as FHIR Validator GUI[30] and Simplifier[31]. The methods section provides a more detailed description of the steps followed for creating the ETL scripts. While building the Python scripts, the mapping of the dictionaries was coded using FHIR-compliant ontologies. The results of the ETL process are JSON files containing the patients’ information standardised to the CDM, one file per patient. These files will serve both as the data source for AI algorithm development, and with proper data requests, standardised data available for the scientific community (Figure 4).

**Figure 4.**
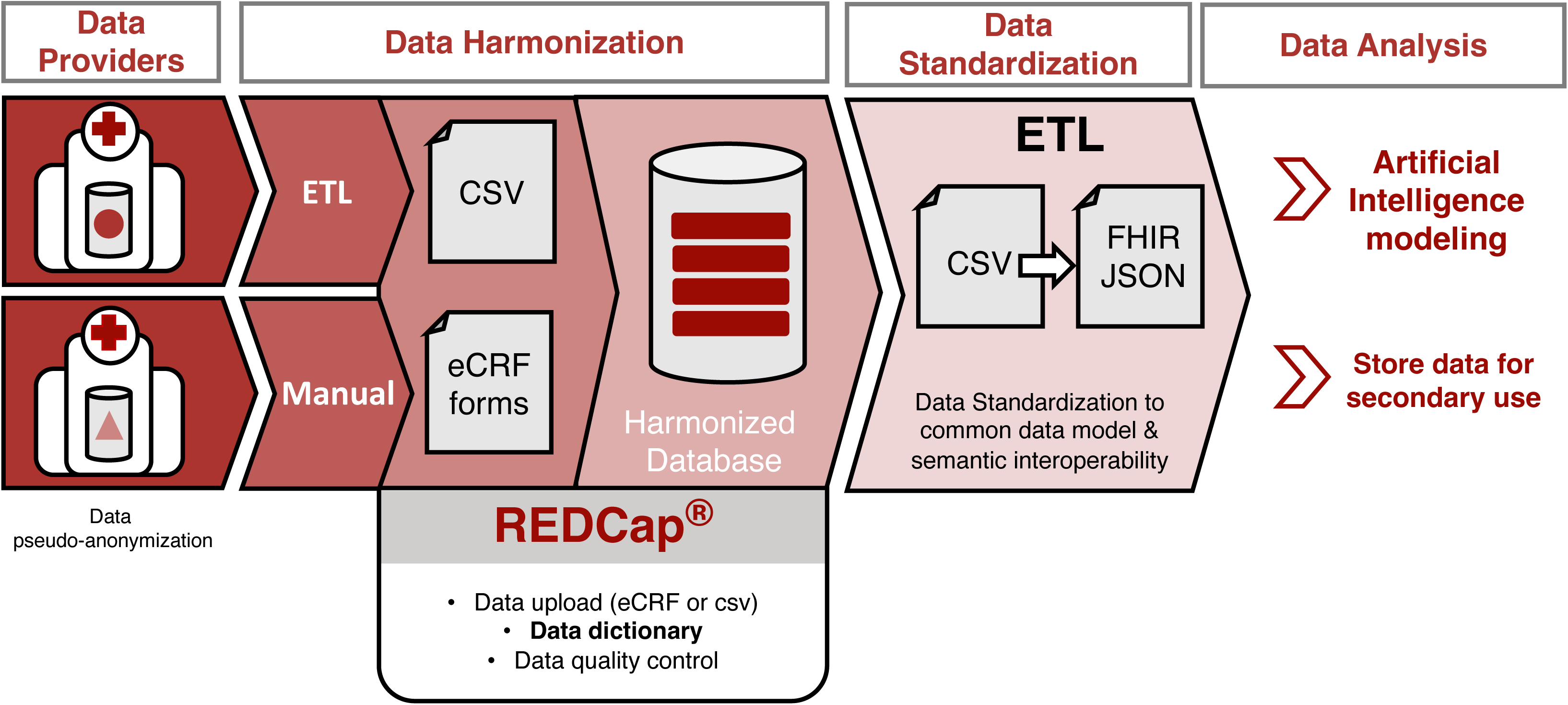
Clinical data processing workflow from clinical institutions to data analysis/sharing. Hospitals import data to REDCap following different paths depending on local resources: 1) some centres introduce data manually (filling online forms or CSV files) or 2) develop their own ETL scripts to automate the process. As a result, all clinical data from the different institutions are merged into a single harmonised database for each use case. Finally, all data is exported from REDCap as a CSV file for standardisation and conversion into FHIR-compliant files (FHIR JSON) and stored at the European Genome-Phenome Archive (EGA).

### Data quality & consistency

Quality data can be defined as data that is fit for purpose, e.g., the data are sufficient for the specified purpose for which it is intended[32,33]. In most cases, data quality for the purpose of machine learning cannot be limited to a single focus but must cater to the needs of multiple audiences. Data quality issues can be introduced at any point in the data management and collection lifecycle. Whether during data acquisition, storage, analysis, or publication, diminished quality can inadvertently affect downstream tasks such as AI training[32–34].

We employ both quality assurance and quality control techniques over the course of the data life cycle, including strict conformance to requirements during input and the assessment methods for repeatable feedback and improvement. The overall objective of our quality analysis is to rate the individual records based on multiple dimensions of quality and use this as a filter for downstream tasks. To achieve the measure of data quality needed for superior AI training and results, we defined quality control rules and procedures based on standard dimensions of quality and built tools to integrate into our pipelines for data collection and storage.

Since we use REDCap as the intermediary data store where all collection methods funnel their data, we found REDCap’s data quality module[27,28] valuable for organising our data quality rules. This module allows for the execution of quality checks for all data entered into the system, whether by direct entry or by CSV imports. This also enabled the capability to export these rules for use in customised tools for data collection.

Data quality can be evaluated over many different dimensions[33] and we have focused our evaluation on three critical dimensions: completeness, conformance and plausibility. For completeness, we focused on value requirements. For conformance, we analysed the various data types and permissible values to ensure adherence. Plausibility applied to ranges, such as age. The types and dimensions are outlined in Table 1.

Much of the needed data quality assessment functionality was already built into the REDCap quality module including pre-established rules handling blank values, data type errors, outliers and invalid permissible values. We also included custom rules covering multiple levels of requirements: minimal, mandatory and recommended. These rules aligned with the required fields outlined for each use case and agreed upon by representatives from each clinical centre.

After assessment of the data based on these organising quality criteria, we generate a score for each quality check based on the number of successes and failures. Here, we can also apply weights if we deem a particular assessment more important than others. Table 2 shows an example of a scoring report.

## LEGAL ASPECTS

In EuCanImage, ethico-legal discussions have played an important role from the beginning of the project and continue to be a recurring topic in similar consortia and initiatives. In addition to the well-known General Data Protection Regulation (GDPR), the last decade has witnessed a surge in regulations and norms that are directly or indirectly relevant when trying to implement data standards for data interoperability. These include, among others, the recently passed Data Governance Act (DGA), the Artificial Intelligence Act (AIA) and the proposal for a Regulation on the European Health Data Space (EHDS). Such regulatory developments are likely to be of considerable practical relevance to the scope of this paper, in particular, for discussions on the personal or non-personal nature of datasets involved, legal barriers for secondary uses of data for AI research and development and cross-border data sharing for AI research in oncology.

### Personal data vs non-personal data

As anticipated, early discussions within research projects and consortia often concern the personal or non-personal nature of the data to be processed. This is mainly due to the fact that processing operations involving personal data, namely information relating to an identified or identifiable natural person, fall under the scope of the GDPR. Therefore, non-personal data such as anonymised data (i.e., personal data rendered anonymous in such a manner that the data subject is not or no longer identifiable) are not bound by the Regulation. Pseudonymised data, which could be attributed to a natural person by the use of additional information, would qualify as personal data (see Article 4(5) and Recital 26 of the GDPR[35]). Generally, de-identification, understood as the process of removing or substituting all personal information and identifiers, is not sufficient to achieve the anonymisation threshold required in the EU. As a result, if the raw data is retained at the source and any key or additional information can be used to reverse the process and re-identify the data subject, this information shall be considered pseudonymised data and thus subject to the GDPR.

Within EuCanImage, all direct and indirect personal identifiers have been removed. In addition, the data is double-hashed both by the data providers and by the platform. Ultimately, all processing operations within the project are aligned with the GDPR and supported by a contractual and governance structure enabling compliant data sharing.

### Secondary use of data for AI research in precision medicine

Further processing of personal data for scientific research purposes, which comprises AI research in precision medicine as pursued by EuCanImage, is compatible with the GDPR (art. 5(1)(b)). Moreover, under the forthcoming EHDS Regulation, secondary use of pseudonymised electronic health data is expressly permitted for “training, testing and evaluating of algorithms, including in medical devices, in vitro diagnostic medical devices, AI systems and digital health applications (art. 34(1)(e)). It should be noted, nonetheless, that these two terms (“further processing” and “secondary use”) are not legally analogous, but the latter will be preferred here for the sake of clarity.

### Cross-border data sharing for AI research in precision medicine

Despite the advent of the GDPR and the harmonisation effort, processing operations involving multiple organisations and researchers from several EU Member states still face slight differences between national, regional and sectoral regulatory frameworks. Hence, although data sharing within Member states of the European Economic Area (EEA) is not hindered by any additional requirements, EuCanImage’s partners and their legal teams still must cope with a complex and fragmented scenario not only from a legal interoperability perspective but also concerning divergent ethical oversight layers and internal procedures at each centre, hospital, institution or country.

Transfers of personal data to third countries or international organisations (i.e., data sharing with researchers or organisations outside the EEA) remain controversial [36,37]. Even though EuCanImage members do not plan to store or process data outside the EEA, controversies have arisen in relation to the transfer of data to international organisations and UK-based institutions after Brexit. Potential routes for transfers remain limited, particularly in light of the strict requirements and threshold set by the Court of Justice of the European Union[38,39].

## DISCUSSION

In many scientific disciplines, especially in health research, working with large scale datasets and engaging in cross-border data sharing is becoming increasingly vital for the adoption and development of AI technologies. EuCanImage focuses on leveraging existing healthcare data to address various scientific research questions using AI models. Our experience uncovers obstacles to data interoperability and reuse, as well as realistic solutions. In this report, we outline our procedures to achieve data interoperability, including a thorough description of the data model design, the standards used, harmonisation efforts, and methodological aspects concerning the practical implementation, along with legal interoperability considerations. The successful development and deployment of our data models and related standards represent a significant milestone, laying the groundwork for future AI applications in cancer healthcare. Furthermore, our work highlights the need for improvements in data collection, annotation, and cross-border dissemination.

We anticipate that our approach and methods will benefit individual institutions and serve as a guide for future large-scale consortia requiring harmonisation and interoperability of cancer-related clinical data for AI and machine learning advancements. Similar efforts have been developed by other consortia[40], including projects within the AI4HI initiative such as CHAIMELEON, ProCancer-I, Incisive and Primarge[14]. These collective endeavours involve meticulous, collaborative, expert-driven analysis, spanning model design, data curation, standards usage, and infrastructure development. The knowledge and experience gained from our combined efforts are crucial in laying the groundwork for future healthcare data standardisation initiatives for AI research across Europe.

Achieving interoperability in healthcare data raises complex issues that need to be addressed. The development of supervised AI models trained for prediction or classification tasks relies on data labelled with ‘ground truth’ classifications. Reaching a consensus on data labelling requires common standard definitions for diagnosis and agreements on the level of data granularity; these are critical factors that affect the reproducibility and quality of the results[41,42]. Cancer diagnosis involves integrating complex criteria based on a variety of disparate data components, such as pathology reports, laboratory results, radiology findings, and advanced molecular and genetic tests. Close collaboration among different medical specialists has enabled the establishment of key principles for data harmonisation: 1) the selection and definition of essential clinical variables to address medical needs, 2) the identification of common data available across all centres and 3) striking a balance between the volume and granularity of the data that can be provided by various hospitals and the optimal information required for AI models (Figure 3).

Within the healthcare-research ecosystem, data sharing remains a barrier. Yet, it is a crucial mechanism for ensuring that high quality data, obtained through exhaustive and expensive processes like defining data labels, harmonisation tasks, and the use of common standards, can be reused by other researchers and, thus, maximise the impact. The FAIR principles provide the framework for such data re-use[43]. Despite progress in adopting interoperability standards, data from different sources still contain discrepancies. To make data fully reusable and reproducible, methods for data cleaning, harmonisation, and standardisation must be transparent[44].

While the presented work demonstrates the feasibility of using HL7 FHIR to achieve interoperability, it also has limitations. FHIR resources were employed for structural interoperability, while SNOMED, LOINC, NCIT, and RxNorm were mainly used for semantic interoperability. By leveraging the comprehensive information model in FHIR, clinical data can be organised hierarchically in a manner that captures its context and remains unambiguous[45]. However, utilising FHIR to build a model for research oncology presents specific constraints and unique requirements for maintaining data interoperability (described previously in the data model section). To maximise the potential of FHIR and encourage broader adoption in the specialised scientific context of AI for precision medicine, alternative FHIR configurations or detailed methodological explanations should be considered to ensure reproducibility. Currently, the main limitation is that the suitability of the models for developing AI algorithms has not yet been validated.

In summary, we demonstrated that large-scale, real-world, multicenter clinical data harmonisation and curation for AI research is feasible through the use or adaptation of common standards. The standardised datasets we will make available at the end of the project, including data from over 20,000 cancer patients, will provide an invaluable resource for investigators to expand the understanding of these complex diseases and open the door for cutting edge translational research beyond the scope of EuCanImage.

## METHODS

### Data model

The clinical data necessary to address each use case was established by interdisciplinary teams, including clinicians and AI experts, considering different key data aspects. Data was arranged following five different data schemas that were compliant with the FHIR (FHIR Release 4B) architecture. The FHIR Resources used were Patient, Condition, Observation, Procedure, Medication Administration and Diagnostic report.

### Ontologies

HL7/FHIR terminology was used in status profiles required by FHIR. SNOMED (SNOMED version International 2022-12-31) was the preferred terminology for general clinical concepts, ICD-O3 (ICD-O3 version 20220429) for histology, LOINC (LOINC version 2.73) for some test observations and RxNorm (RxNorm version 03-Jan-2023) for medication. We used NCIT (NCIT version 23.8d) when the concept did not exist in previous ontologies.

### Data capture, standardisation and quality control

Patient IDs were pseudo-anonymised at the hospitals, and only hashed patient IDs (EuCanImage ID) were introduced in the platform. Data from hospitals was captured in REDCap [REDCap version 13.10.0; PHP 8.1.3 (Linux/Unix OS); MySQL 8.0.30] by filling the online forms or uploading CSV files complying with the specific format requirements. As a result, all clinical data were merged into a single harmonised database for each data schema. At this stage, we performed quality control checks. We focused our evaluation on three critical dimensions: completeness, conformance and plausibility, and generated a score for each quality check based on the number of successes and failures. We built ETL pipelines in python to transform the harmonised output data into JSON files compliant with FHIR. We used FHIR Validator GUI[30] and Simplifier[31] as external validators for quality control. Code availability: The python scripts to transform harmonised data to the FHIR compatible schemas are available on Github (see ‘Availability of supporting source code and requirements’ section).

### Steps to create the ETL script

**Step 0 - Dictionary:** Before processing the data, we set up an environment with the necessary resources. This includes the creation of a machine readable data dictionary encoding a) the name of the variable, b) the ontological code and c) the RedCap internal codification. This mapping served two purposes simultaneously: 1) Data Quality Control and 2) the Extract Transform Load (ETL) process.
**Step 1 - Parsing:** All the data gathered in REDCap was exported into a CSV file per use case and clinical centre. These CSV files were parsed into a Python readable form for the posterior transformation into the objects required by the libraries.
**Step 2 - Dictionary import:** For the transformations to occur with minimal errors, dictionaries from step 0 were imported into the Python script for the consecutive mapping to the FHIR Resources.
**Step 3 - FHIR Resource mapping:** To streamline the transformation of the different types of data variables into their respective FHIR Resources, we defined functions to automate the process.

1. First, empty templates were created for each FHIR Resource type. To avoid errors, we maintained libraries that followed FHIR structures with internal validators.
2. Then, depending on the input required by each function (such as information about medication administrations, quantitative or qualitative observations, conditions from the patients or others), the Resource was populated accordingly. Additionally, some variables needed extra processing, such as date and timestamp parsing, which is also automated by the code.
3. In the case of an error in the structure, the libraries flag them for correction.
**Step 4 - Export and validation:** As a final step, the objects that were created in the script needed to be exported into JSON files. Since the code included FHIR libraries streamlining the process, parsing the generated objects and dictionaries into the JSON file was straightforward. The libraries used in the process validate the integrity of the structure but not always of the contents. To confirm the correctness of the result, we used external validators, such as FHIR Validator GUI[30] and Simplifier[31].

### European Genome-Phenome Archive

The European Genome-phenome Archive (EGA) is a service for permanent archiving and sharing of personally identifiable genetic, phenotypic, and clinical data. The standardised clinical data, one JSON file (FHIR compliant) per patient obtained after the previously described process, will be encrypted and stored at the EGA repository.

## Data Availability

This study describes a new process to harmonize and standardize clinical data. The data will be available at EGA upon request to the authors.

## DATA AVAILABILITY

An archival copy of the code is available via Software Heritage [46] and Workflowhub [47]. Since EuCanImage is still ongoing the data is not available yet. The datasets will be available at the end of the project under controlled access at the European Genome-Phenome Archive (clinical data) and EuroBioimaging (images).

## AVAILABILITY OF SUPPORTING SOURCE CODE AND REQUIREMENTS

Project name: EuCanImage FHIR implementation

Project home page: https://github.com/EGA-archive/EuCanImage-FHIR/

Operating system(s): Platform independent

Programming language: Python

Other requirements: Python 3.11.2 or higher, FHIR Resources 6.5.0 or higher, pandas 2.1.3 or higher, numpy 1.26.2 or higher

License: Apache License 2.0

RRID: SCR_025824

Workflowhub - EuCanImage FHIR ETL implementation (DOI):

Hepatocellular Carcinoma [10.48546/workflowhub.workflow.1112.1]
Colorectal liver metastasis [10.48546/workflowhub.workflow.1156.1]
Rectal cancer [10.48546/workflowhub.workflow.1157.1]
Breast cancer MMG [10.48546/workflowhub.workflow.1158.1]
Breast cancer MRI [10.48546/workflowhub.workflow.1159.1]

## ACKNOWLEDGEMENTS

We acknowledge the support of the Spanish Ministry of Science and Innovation through the Centro de Excelencia Severo Ochoa (CEX2020-001049-S, MCIN/AEI /10.13039/501100011033) and the Generalitat de Catalunya through the CERCA programme. We are grateful to the CRG Core Technologies Programme for their support and assistance in this work.

We acknowledge the Social and Legal Sciences Applied to the New Technosciences Research Group, University of the Basque Country (UPV/EHU). Grant from the Department of Education of the Basque Government to support the activities of Research Groups from the Basque University System (Reference IT 1541-22)

This project has received funding from the European Union’s Horizon 2020 research and innovation programme under grant agreement No 952103.

## AUTHOR CONTRIBUTIONS

J.R., L.F. and T.G.L. developed the concept. F.P., K.L., M.S., M.B., and S.C. substantially contributed to the development of the concept. M.B., K.R., S.F., E.N. provided clinical advice. J.R., L.F., T.G.L., M.B., K.R., S.F. and R.F. conceptualised the data model. A.C. and M.Ry. Implemented the model and tests. A.C. and M.Ru. wrote the code. M.Ru., F.P., J.B. and A.C. conceptualised and implemented quality control. M.Re. and P.L. provide legal guidelines. T.G.L., M.B., S.F., M.Ru., M.Re., A.C. and AN conceptualised and wrote the manuscript. T.G.L. and M.B. created figures. All authors contributed to the critical revision of the manuscript.

## COMPETING INTERESTS

The authors declare no competing interests

EuCanImage CONSORTIUM (Lead authors extended list)

**Jordi Rambla -** Centre for Genomic Regulation (CRG), The Barcelona Institute of Science and Technology, Dr.Aiguader 88, Barcelona 08003, Spain

**Maciej Bobowicz** - 2nd Department of Radiology, Medical University of Gdansk, Mariana Smoluchowskiego 17, 80-214, Gdansk, Poland

**Jordi Rimola &** Xavier Bargalló - Fundació Clínic per a la recerca biomedica, Carrer Rossello 149, 08036 Barcelona, Spain

**Fred Prior** - Department of Biomedical Informatics, University of Arkansas for Medical Sciences, Little Rock, Arkansas, United States

**Pilar Nicolas** - Social and Legal Sciences Applied to the New Technosciences Research Group, University of the Basque Country (UPV/EHU), Bilbao, Spain

**Katrine Riklund** - Department of DIagnostics and Intervention, Diagnostic Radiology, Umeå university, Umeå, Sweden

**Lorenzo Faggioni -** Academic Radiology, Department of Translational Research, University of Pisa, Via Roma 67, 56126 Pisa, Italy

**Josep Lluis Gelpi** - Barcelona Supercomputing Center (BSC), Jordi Girona 29, 08034, Barcelona, Spain.

**Martijn P. A. Starmans** - Department of Radiology and Nuclear Medicine and Department of Pathology, Erasmus MC Medical Center, Rotterdam, the Netherlands

**Kaisar Kushibar & Karim Lekadir** - Artificial Intelligence in Medicine Lab (BCN-AIM), Departament de Matemàtiques i Informàtica, Universitat de Barcelona, Barcelona, Spain

**Karim Lekadir** - Institució Catalana de Recerca i Estudis Avançats (ICREA), Passeig Lluís Companys 23, Barcelona, Spain.

**Philippe Lambin & Henry Woodruff** - Universiteit Maastricht (UM) Minderbroedersberg 4- 6, 6211 LK Maastricht, The Netherlands

**Melanie Goisauf** - Biobanks and Biomolecular Resources Research Infrastructure Consortium (BBMRI-ERIC), Neue Stiftingtalstrasse 2/B/6, 8010 Graz, Austria

**Davide Zaccagnini** - Lynkeus (LYN), Livenza 6, 00198 Rome, Italy

**Pär Kragsterman** - Collective Minds Radiology AB (CMRAD), Holrnalkersvalgen 14, 183 65 Talby, Sweden

**Flore Belmans** - Oncoradiomics (ONCO), Clos Chanmurly 13, 4000 Liege, Belgium

**Tobias Heimann** - Siemens Healthcare GMBH (SIE), Henkestr.127, 91052 Erlangen, Germany

**Peter Gordebeke** - EIBIR Gemeinnutzige GMBH zur Forderung der Erforschung der Biomedizinischen Bildgebung (EIBIR), Neutorgassse 9/2, 1010 Wien, Austria

**Emanuele Neri** - European Society of Oncologic Imaging - ESOI Europaische Gesellschaftfur Onkologische Bildebung (ESOI), Am Gestade 1, 1010 Vienna, Austria

**Jane Smith** - European Association for Cancer Research (EACR), Sir Colin Campbell Building, Triumph Road, Nottingham, NG72TU, UK

**Juozas Kup**č**inskas** - Lietuvos Sveikatos Mokslu Universiteto Ligonine KaunoKlinikos (KAUNO), Eiveniu Street 2, LT-50161 Kaunas, Lithuania

**Javier del Riego** - Institut d’Investigació i Innovació Parc Taulí I3PT. Universitat Autònoma de Barcelona. Sabadell, Barcelona, Spain

## REFERENCES

1. Lotter W, Hassett MJ, Schultz N, Kehl KL, Van Allen EM, Cerami E. Artificial Intelligence in Oncology: Current Landscape, Challenges, and Future Directions. Cancer Discov. 2024; doi: 10.1158/2159-8290.CD-23-1199.

2. Haendel MA, Chute CG, Robinson PN. Classification, Ontology, and Precision Medicine. N Engl J Med. 2018; doi: 10.1056/NEJMra1615014.

3. Dinov ID. Methodological challenges and analytic opportunities for modeling and interpreting Big Healthcare Data. GigaScience. 2016; doi: 10.1186/s13742-016-0117-6.

4. Wilkinson MD, Dumontier M, Aalbersberg IjJ, Appleton G, Axton M, Baak A, et al.. The FAIR Guiding Principles for scientific data management and stewardship. Sci Data. Nature Publishing Group; 2016; doi: 10.1038/sdata.2016.18.

5. Vesteghem C, Brøndum RF, Sønderkær M, Sommer M, Schmitz A, Bødker JS, et al.. Implementing the FAIR Data Principles in precision oncology: review of supporting initiatives. Brief Bioinform. 2020; doi: 10.1093/bib/bbz044.

6. He J, Baxter SL, Xu J, Xu J, Zhou X, Zhang K. The practical implementation of artificial intelligence technologies in medicine. Nat Med. 2019; doi: 10.1038/s41591-018-0307-0.

7. Kruse CS, Goswamy R, Raval YJ, Marawi S. Challenges and Opportunities of Big Data in Health Care: A Systematic Review. JMIR Med Inform. 2016; doi: 10.2196/medinform.5359.

8. Sweeney SM, Hamadeh HK, Abrams N, Adam SJ, Brenner S, Connors DE, et al.. Challenges to Using Big Data in Cancer. Cancer Res. 2023; doi: 10.1158/0008-5472.CAN-22-1274.

9. Frid S, Bracons Cucó G, Gil Rojas J, López-Rueda A, Pastor Duran X, Martínez-Sáez O, et al.. Evaluation of OMOP CDM, i2b2 and ICGC ARGO for supporting data harmonization in a breast cancer use case of a multicentric European AI project. J Biomed Inform. 2023; doi: 10.1016/j.jbi.2023.104505.

10. Näher A-F, Vorisek CN, Klopfenstein SAI, Lehne M, Thun S, Alsalamah S, et al.. Secondary data for global health digitalisation. Lancet Digit Health. 2023; doi: 10.1016/S2589-7500(22)00195-9.

11. Ayaz M, Pasha MF, Alzahrani MY, Budiarto R, Stiawan D. The Fast Health Interoperability Resources (FHIR) Standard: Systematic Literature Review of Implementations, Applications, Challenges and Opportunities. JMIR Med Inform. 2021; doi: 10.2196/21929.

12. Kondylakis H, Kalokyri V, Sfakianakis S, Marias K, Tsiknakis M, Jimenez-Pastor A, et al.. Data infrastructures for AI in medical imaging: a report on the experiences of five EU projects. Eur Radiol Exp. 2023; doi: 10.1186/s41747-023-00336-x.

13. Marti-Bonmati L, Koh D-M, Riklund K, Bobowicz M, Roussakis Y, Vilanova JC, et al.. Considerations for artificial intelligence clinical impact in oncologic imaging: an AI4HI position paper. Insights Imaging. 2022; doi: 10.1186/s13244-022-01220-9.

14. Kondylakis H, Ciarrocchi E, Cerda-Alberich L, Chouvarda I, Fromont LA, Garcia-Aznar JM, et al.. Position of the AI for Health Imaging (AI4HI) network on metadata models for imaging biobanks. Eur Radiol Exp. 2022; doi: 10.1186/s41747-022-00281-1.

15. SNOMED International: The SNOMED CT browser. https://www.snomed.org Accessed 2024 Feb 1.

16. World Health Organization: International Classification of Diseases (ICD-11). https://www.who.int/standards/classifications/classification-of-diseases Accessed 2024 Feb 1.

17. LOINC: SearchLOINC. https://loinc.org/ Accessed 2024 Feb 5.

18. Reich C, Ostropolets A, Ryan P, Rijnbeek P, Schuemie M, Davydov A, et al.. OHDSI Standardized Vocabularies-a large-scale centralized reference ontology for international data harmonization. J Am Med Inform Assoc JAMIA. 2024; doi: 10.1093/jamia/ocad247.

19. International Cancer Genome Consortium: The ICGC ARGO data dictionary. https://www.icgc-argo.org/ Accessed 2023 Oct 5.

20. Health Level Seven International (HL7): Fast Healthcare Interoperability Resources (FHIR). https://www.hl7.org/fhir/ Accessed 2023 Aug 30.

21. Beale SH Thomas: openEHR International. https://www.openehr.org/ Accessed 2024 Feb 1.

22. Plazzotta F, Luna D, González Bernaldo de Quirós F. [Health information systems: integrating clinical data in different scenarios and users]. Rev Peru Med Exp Salud Publica. 32:343–512015;

23. Garza M, Del Fiol G, Tenenbaum J, Walden A, Zozus MN. Evaluating common data models for use with a longitudinal community registry. J Biomed Inform. 2016; doi: 10.1016/j.jbi.2016.10.016.

24. Duda SN, Kennedy N, Conway D, Cheng AC, Nguyen V, Zayas-Cabán T, et al.. HL7 FHIR-based tools and initiatives to support clinical research: a scoping review. J Am Med Inform Assoc JAMIA. 2022; doi: 10.1093/jamia/ocac105.

25. Vorisek CN, Lehne M, Klopfenstein SAI, Mayer PJ, Bartschke A, Haese T, et al.. Fast Healthcare Interoperability Resources (FHIR) for Interoperability in Health Research: Systematic Review. JMIR Med Inform. 2022; doi: 10.2196/35724.

26. Observational Health Data Sciences and Informatics: The OMOP Common Data Model. https://ohdsi.github.io/CommonDataModel/index.html Accessed 2023 Oct 5.

27. Harris PA, Taylor R, Minor BL, Elliott V, Fernandez M, O’Neal L, et al.. The REDCap consortium: Building an international community of software platform partners. J Biomed Inform. 2019; doi: 10.1016/j.jbi.2019.103208.

28. Harris PA, Taylor R, Thielke R, Payne J, Gonzalez N, Conde JG. Research electronic data capture (REDCap)—A metadata-driven methodology and workflow process for providing translational research informatics support. J Biomed Inform. 2009; doi: 10.1016/j.jbi.2008.08.010.

29. Cheng AC, Duda SN, Taylor R, Delacqua F, Lewis AA, Bosler T, et al.. REDCap on FHIR: Clinical Data Interoperability Services. J Biomed Inform. 2021; doi: 10.1016/j.jbi.2021.103871.

30. HL7 FHIR: Validator-wrapper. https://validator.fhir.org/ Accessed 2024 Sep 2.

31. HL7-FHIR: SIMPLIFIER.NET. https://simplifier.net/ Accessed 2024 Sep 2.

32. Talburt JR. Entity Resolution and Information Quality. Elsevier;

33. Bian J, Lyu T, Loiacono A, Viramontes TM, Lipori G, Guo Y, et al.. Assessing the practice of data quality evaluation in a national clinical data research network through a systematic scoping review in the era of real-world data. J Am Med Inform Assoc JAMIA. 2020; doi: 10.1093/jamia/ocaa245.

34. Priestley M, O’donnell F, Simperl E. A Survey of Data Quality Requirements That Matter in ML Development Pipelines. J Data Inf Qual. 2023; doi: 10.1145/3592616.

35. Regulation (EU) 2016/679 of the European Parliament and of the Council of 27 April 2016 on the protection of natural persons with regard to the processing of personal data and on the free movement of such data, and repealing Directive 95/46/EC (General Data Protection Regulation). 2016 May 4;

36. Bernier A, Molnár-Gábor F, Knoppers BM, Borry P, Cesar PMDG, Devriendt T, et al.. Reconciling the biomedical data commons and the GDPR: three lessons from the EUCAN ELSI collaboratory. Eur J Hum Genet. Nature Publishing Group; 2024; doi: 10.1038/s41431-023-01403-y.

37. Molnár-Gábor F, Beauvais MJS, Bernier A, Jimenez MPN, Recuero M, Knoppers BM. Bridging the European Data Sharing Divide in Genomic Science. J Med Internet Res. 2022; doi: 10.2196/37236.

38. Court of Justice of the European Union. Judgement of 16 July 2020, Case Schrems II (C-311/18).

39. European Data Protection Board (EDPB). Recommendations 01/2020 on measures that supplement transfer tools to ensure compliance with the EU level of protection of personal data, adopted on 18 June 2021.

40. Sweeney SM, Hamadeh HK, Abrams N, Adam SJ, Brenner S, Connors DE, et al.. Case Studies for Overcoming Challenges in Using Big Data in Cancer. Cancer Res. 2023; doi: 10.1158/0008-5472.CAN-22-1277.

41. Wang SY, Pershing S, Lee AY, Committee on behalf of the AT on A and AMIT. Big data requirements for artificial intelligence. Curr Opin Ophthalmol. 2020; doi: 10.1097/ICU.0000000000000676.

42. Cirillo D, Núñez-Carpintero I, Valencia A. Artificial intelligence in cancer research: learning at different levels of data granularity. Mol Oncol. 2021; doi: 10.1002/1878-0261.12920.

43. Scheffler M, Aeschlimann M, Albrecht M, Bereau T, Bungartz H-J, Felser C, et al.. FAIR data enabling new horizons for materials research. Nature. 2022; doi: 10.1038/s41586-022-04501-x.

44. Heacock ML, Lopez AR, Amolegbe SM, Carlin DJ, Henry HF, Trottier BA, et al.. Enhancing Data Integration, Interoperability, and Reuse to Address Complex and Emerging Environmental Health Problems. Environ Sci Technol. American Chemical Society; 2022; doi: 10.1021/acs.est.1c08383.

45. Leroux H, Metke-Jimenez A, Lawley MJ. Towards achieving semantic interoperability of clinical study data with FHIR. J Biomed Semant. 2017; doi: 10.1186/s13326-017-0148-7.

46. EuCanImage-FHIR [Software Heritage]. 2024. https://urldefense.com/v3/_https://urldefense.com/v3/https://archive.softwareheritage.org/swh:1:dir:665885f643873d80daa240661e8c2bc6736bd47c;origin=https:/*github.com/EGA-archive/EuCanImage-FHIR;visit=swh:1:snp:884dce6e07416fad00a2eb80e2306220bf28d8fa;anchor=swh:1:rev:17d46f996865c538354a8ead84e171028208b194;Lw!!D9dNQwwGXtA!SuPF6HZiw5PJbZ0HJD5N-hucLshPU5ORQTjEWsiAS9DPeB_be8NRdgp6OZFYb9O_VhfNbnmXi0nCH9bTKgI$

47. EuCanImage FHIR ETL implementation. 2024. https://workflowhub.eu/collections/25

